# Spatial and Bulk Transcriptomics Reveal Distinct Molecular Signatures in Kaposi Sarcoma with and Without other KSHV-Associated Diseases

**DOI:** 10.64898/2025.11.30.25341287

**Authors:** Quashawn Chadwick, Ned Cauley, Jose Mercado-Matos, Bahman Afsari, Xiaolin Wu, Laura Bassel, Maria Hernandez, Michelly Sampaio De Melo, Xiaofan Li, Kathryn Lurain, Robert Yarchoan, Joseph Ziegelbauer, Christopher A. Febres-Aldana, Laurie T. Krug, Ramya Ramaswami

## Abstract

**Background:** Kaposi sarcoma (KS), caused by Kaposi sarcoma herpesvirus (KSHV), is an angioproliferative tumor that presents as skin lesions in people with HIV. Other KSHV-associated diseases (KAD) including multicentric Castleman disease (MCD), primary effusion lymphoma (PEL), and KSHV-associated inflammatory cytokine syndrome (KICS) may occur concurrently with KS and influence clinical outcomes. New sequencing technologies enabling analysis of archival KS tissues provide mechanisms to define cellular and viral characteristics that contribute to disease heterogeneity.

**Methods:** We profiled gene expression in 42 confirmed KS formalin-fixed paraffin-embedded (FFPE) skin biopsies using the Nanostring nCounter PanCancer ImmunoOncology panel supplemented with KSHV-specific probes. Spatial RNA profiling was performed on four tissues from participants with KS and concurrent KAD (KS+KAD) using GeoMx digital spatial profiling (DSP) platform. Regions of interest were selected using LANA-1, CD45 and CD31 staining to characterize tumor (LANA-1^+^, CD31^+^), vessel (LANA-1, CD31^+^) and immune cells (CD45^+^) areas of illumination (AOIs).

**Results:** KS samples were obtained from 42 men with HIV (median age 40 years). Median HIV viral load of 27 copies/mL and median CD4^+^ T-cell count was 211 cells/µL. Forty-eight percent had KS alone and 52% had KS+KAD. Patients with KS+KAD had worse survival compared to those with KS alone. Transcriptomic analyses identified increased expression of STC1 (log2FC=2.02, p-adjusted (padj)=0.001), a secreted glycoprotein, and MKI67 (log2FC=1.11, padj=0.02), a common proliferation marker, in KS+KAD lesions, along with lower expression of cytokine-associated pathways. Spatial RNA profiling from 4 KS samples from patients with KS+KAD identified increased abundance of lymphatic endothelial cells, elevated LYVE1 expression in LANA-1+ tumor areas as compared to LANA- areas.

**Conclusions:** Bulk and spatial transcriptomic profiling of archival HIV-associated KS lesions revealed disease-specific molecular programs associated with concurrent KAD that altered tumor and microenvironment features. These findings demonstrate the heterogeneity of KS lesions that may guide future studies on KS pathogenesis and potential therapeutic targets.

## Introduction

Kaposi sarcoma herpesvirus (KSHV) is an oncogenic virus with a seroprevalence of 3-7% in the general population of the United States(1, 2). It has a substantially higher seroprevalence of 38-70% in the population of men who have sex with men (MSM) (3-5). KSHV causes Kaposi sarcoma (KS)(6), an angio-proliferative tumor associated with KSHV infection of endothelial cells that typically manifests as hyperpigmented skin lesions but can also affect other organs, including the pulmonary and gastrointestinal (GI) tracts (7) in advanced cases. Within the United States, epidemic KS is the most common form, which is observed among people with HIV (PWH) at any CD4^+^ T-cell count(8). KS remains one of the most common cancers in areas of Uganda, Malawi and other countries in sub-Saharan Africa, where KSHV is endemic and HIV co-infection can impact morbidity and mortality (9, 10). The diagnosis of KS requires histologic confirmation from affected tissue, demonstrating spindle cells positive for KSHV latency-associated nuclear antigen (LANA-1) and endothelial markers (CD31+ and CD34+) on immunohistochemistry staining.

In addition to KS, KSHV is also the causative agent of other KSHV-associated diseases (KAD). This includes multicentric Castleman disease (MCD)(11), a lymphoproliferative disorder and primary effusion lymphoma (PEL)(12), a type of B-cell non-Hodgkin lymphoma. KSHV-associated inflammatory cytokine syndrome (KICS) is associated with KSHV viremia and systemic inflammatory signs and symptoms. KICS requires the exclusion of PEL and MCD (13). MCD, PEL and KICS are characterized by elevated inflammatory cytokines in the circulation, such as interleukin (IL)-6 and IL-10(14-17). KSHV also encodes a viral homolog of IL-6 (v-IL6), which has been associated with angiogenesis and tumorigenesis in KAD(18, 19). KS can occur concurrently with KAD, particularly among PWH and these disorders can be challenging to diagnose as they require sampling of lymph nodes and/or effusions in patients who may be critically ill(15, 20, 21).

The extent of KS involvement and the presence of concurrent KAD impact the overall management and outcomes. Among PWH, antiretroviral therapy (ART) is an essential component of KS therapy. In those with symptomatic or extensive disease, chemotherapy and immunomodulatory agents are standard treamtnets used in addition to ART(22). When KS occurs with KAD, treatment should be directed at any additional diagnoses (PEL and/or MCD, or KICS)(20). Although KS may occur alone or with KAD, there is limited understanding of differences in gene expression profiles of KS tissue in the context of KAD.

Previous studies evaluating KS using bulk RNA sequencing methods have identified several key genes and pathways that impact KS pathogenesis. A previous study of 4 PWH and cutaneous KS identified altered lipid and glucose metabolism and higher levels of B cells, macrophages and NK cells in KS tissues as compared to controls(23). Evaluation of 22 cutaneous and GI KS lesions as compared to normal paired tissue identified higher IL-6 and IL-10 gene expression in cutaneous KS lesions but this was not observed in GI KS lesions(24). This study also identified 26 cellular genes that were highly expressed in KS lesions as compared to normal tissue. This included *STC1*, which encodes a secreted glycoprotein, and *FLT4*, encoding a tyrosine kinase receptor for vascular endothelial growth factors (25, 26). Knockdown of *STC1* and *FLT4* impaired tubule formation in KSHV-infected lymphatic endothelial cells. Therefore, identifying differentially expressed genes in patients with distinct clinical characteristics may provide insights into KS pathogenesis and potential therapeutic targets for clinical trials.

Despite the relatively uniform descriptions of KS histopathology, patients with KS exhibit significant heterogeneity in clinical characteristics. Factors such as immune microenvironment, CD4+ T-cell count, the presence of concurrent KAD are commonly associated with cytokine dysregulation and influence patient outcomes (15, 27, 28). However, it is unclear whether these characteristics are associated with distinct gene expression profiles, which could aid subtyping in KS lesions. Here, we used the nCounter gene expression assay and spatial transcriptomic analysis to evaluate phenotypic and disease-specific differences in transcriptomic profiling of archival cutaneous KS lesions.

## Methods

### Patient cohort and specimen collection

Archived samples from patients with KS under the care of the HIV/AIDS Malignancy Branch at the National Cancer Institute who had consented to sequencing of their tissue were included in the analyses (Figure 1A). HIV characteristics and clinical demographics, including KAD type and prior treatment, were obtained at the time of biopsy collection. Those included in these analyses had KS biopsies obtained and provided consent to the protocols for sample collection and genomic analyses. KSHV viral load (VL) in peripheral blood mononuclear cells (PBMCs) was assessed by quantitative real-time polymerase chain reaction as previously described (29).

**Figure 1.**
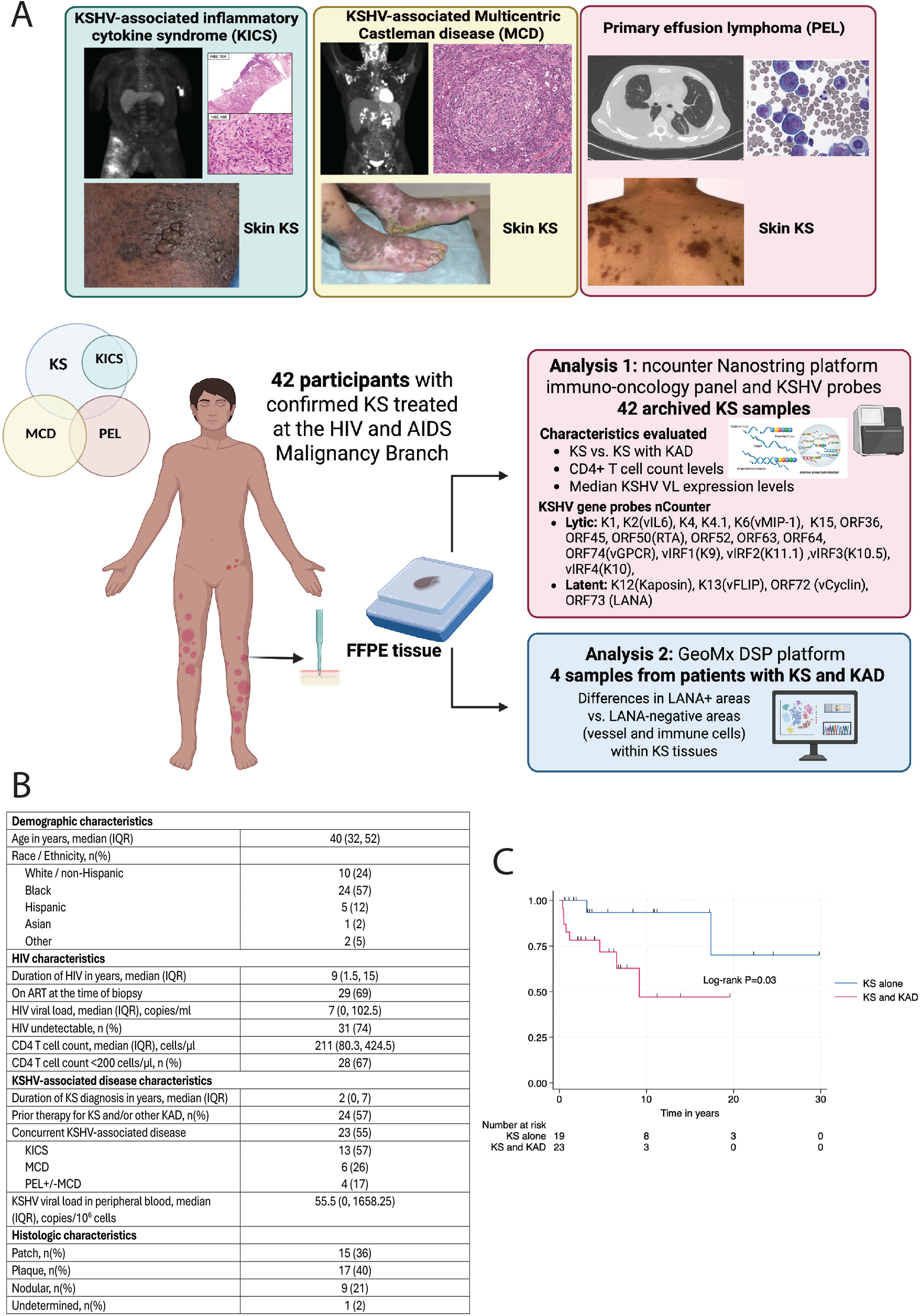
Overall workflow and gene expression profile differences between patients with KS and those with KS and other KAD. A. Summary of workflow and analyses of KS lesion specimens B. Distribution of age, HIV viral load, CD4 T-cell count, and prevalence of concurrent KSHV-associated diseases (KAD) among 42 individuals with HIV and cutaneous Kaposi sarcoma. Median values and interquartile ranges are shown. C. Kaplan Meier curve of overall survival stratified by concurrent KS and KAD diagnosis.

All participants consented to protocols for tissue procurement (NCT00006518) and genomic sequencing of KS and other KSHV-associated diseases (NCT03300830). Both protocols were approved by the NIH Institutional Review Board. All enrolled participants gave written informed consent in accordance with the Declaration of Helsinki.

### nCounter Analysis

RNA was extracted from FFPE samples and run on the nCounter Analysis System (NanoString Technologies https://nanostring.com/products/ncounter-analysis-system/ncounter-systems-overview/), according to the manufacturer’s protocol. All hybridizations were 17-22 hours long, and all counts were gathered by scanning on HIGH mode for 280 fields of view per sample. Quality control (QC) checks were applied to raw count files for each sample according to the nCounter Gene Expression Analysis Guidelines(30), and applied in R Studio using R functions adapted from Battacharya et al(31). After manual inspection of all samples flagged from QC checks and evaluated with principal component analysis (PCA) plots, no samples were removed for downstream analysis. Housekeeping genes provided in the probe set were evaluated for differential expression between sample groups using a general linear model and no housekeeping genes were found to be significantly different. Upper-quartile normalization was applied to correct for technical differences between samples. Unaccounted variation after upper quartile normalization was determined using RUVseq with 1 factor of variation using the probe set’s housekeeping genes(32). For each differential expression comparison, the factor generated by RUVseq was added as a covariate using the DESeq function of DESeq2(33).

Read counts were log-transformed for selected cytokine comparisons between patients with KS and those with other KAD. To understand differences in KSHV expression within each sample, the median expression of latent and lytic genes was obtained. These values were log-transformed and compared between those with KS versus KS and concurrent KAD and by CD4 T-cell count (<200 cells/µl (Low) vs ≥200cells/µl (High)) using Wilcoxon signed-rank test.

We determined overall survival differences between those with KS as compared to patients with KS and other concurrent KAD using the Kaplan-Meier method and log-rank test. We investigated whether the nCounter gene expression were predictive of survival outcomes in those with KS. A Cox Model directly to the continuous gene expression, after applying a log2 transformation to the RUV-normalized gene expression, and a forest plot was analyzed for the hazard ratio for genes with a P-value < 0.05. Due to the exploratory nature and small sample size of these survival analyses, the P-values were unadjusted.

### DSP Analysis

Four FFPE tissue samples from patients with concurrent KAD (2 with MCD and KS and 2 with KICS and KS) were processed for GeoMx whole-transcriptome analysis following the manufacturer^’^s recommendations (NanoString/Bruker). Briefly, 5 µm sections were deparaffinized and subjected to antigen retrieval (ER2, 100^°^C for 20 min). Sections were fixed in neutral-buffered formalin (NBF), treated with proteinase K (1 µg^·^mL^−1^; Ambion, Austin, TX) on a Leica Bond RX autostainer, and hybridized overnight with the WTA probe set at 37^°^C in a hybridization oven. After stringent washes (50% formamide in 2^×^ SSC) and morphology staining, slides were counterstained with SYTO 83 (nuclei), stained for KSHV LANA-1 (LN53, Abcam, ab4103; donkey anti-rat AF-647, Thermo Fisher A48272) to identify infected cells, and for endothelial markers (CD31, Bio-Techne AF3628; donkey anti-goat AF-488, Thermo Fisher A-11055). Staining pattern and specificity were reviewed by a board-certified pathologist.

Fluorescent whole-slide images were scanned, and regions of interest (ROIs)^—^rectangular, circular, or free-hand polygons^—^were selected based on histology and marker expression to capture representative KS tumor, vascular, and immune regions. Within each ROI, areas of illumination (AOIs) were defined using marker-based segmentation or manual selection to capture discrete morphological compartments. Uninfected vessel AOIs were defined by regions with CD31^+^LANA-1^+^ staining and collected as full polygons. Tumor AOIs were identified as CD31^+^LANA-1^+^ regions, and samples were extracted either from the entire AOI or restricted to LANA-1^+^ nuclear regions within the AOI.

Photocleavable oligonucleotide barcodes from each AOI were released by targeted UV illumination, collected into 96-well plates, and processed for next-generation sequencing according to the GeoMx NGS protocol. Libraries were sequenced on a NextSeq 2000 (P3 flow cell; 27 ^×^ 27 bp paired reads with 8 × 8 index reads). Basecall files were demultiplexed using bclconvert v3.8.4, and FASTQs were processed to Digital Count Conversion (DCC) files using the GeoMx NGS Pipeline v2.3.3.10.

DSP analysis was performed using the R Package DSP Workflow(34). Raw count files were combined with annotations and probes were mapped to gene names using the Nanostring R package GeoMxTools(35). Raw count files were combined with annotations and probes were mapped to gene names for the human WTA probe set. QC checks were applied in R studio using Nanostring’s guidelines for best practices of analysis of DSP data from RNA probes with NGS sequencing(36). Areas of illumination (AOIs) and probes that were flagged from QC checks or with less than 1% detection were removed before downstream analysis. Upper-quartile normalization was applied after QC. PCA plots were used to evaluate the effect of normalization and to check for batch effects. Differential expression analysis was performed using GeoMxTools’s mixed model function with the slide number used as a random intercept, and a random slope was added when comparing AOIs within the same slide(37).

Gene Set Enrichment (GSEA) analysis was ran using the R package ClusterProfiler(38). Genes for each differential expression comparison output were ranked according to the Signal2Noise score, which were calculated using the formula defined in the GSEA manual(39).

Cell type deconvolution was performed using the R package SpatialDecon(40). The references “Skin_HCA” and “ImmuneCensus_HCA” were used according to the package instructions. All plots were generated with custom R functions using the R package ggplot2(41). The R package ggpubr was used for running Wilcoxon rank sum tests on cell type abundance scores between annotation groups and displaying significance in boxplots(42). Gene functions were determined using the National Library of Medicine Center for Biotechnology Information Gene Reference Database.

## Results

### Patient characteristics and differences in KS and KAD survival outcomes

Archival cutaneous KS samples were obtained from 42 men with HIV with a median age of 40 years (interquartile range (IQR): 32-52 years, **Figure 1B**). Fifty-seven percent of the patients were Black. The median duration of HIV infection was 9 years (IQR: 1.5-15 years), and patients had a median HIV viral load of 27 copies/mL (IQR: 0 – 103 copies/mL). The median CD4+ T-cell count was 211 cells/µL (IQR: 80 – 425 cells/µL). Twenty-three (55%) of the patients had KS and other concurrent KAD. Among those with concurrent KS and KAD, the most common diagnosis was KICS with KS, observed in 12 (55%) participants. An additional 6 (27%) participants had MCD and KS, 2 (9%) participants had PEL and KS, and 2 (9%) participants had MCD, PEL and KS. With regard to histologic stage, the majority of the analyzed specimens were KS lesions at the patch or plaque stage, and 21% were nodular.

Among patients with KS in this cohort, a concurrent KAD diagnosis was associated with poor outcome as compared to having KS alone (10-year overall survival for KS alone was 92% vs. 47% for those with KS and other concurrent KAD, log-rank test P=0.03, **Figure 1C**). Given the survival differences noted between these characteristics, our evaluation of these KS tissues focused on comparing KS with KS concurrent with other KAD (KS+KAD).

### Differential gene expression within KS lesions varies by the presence of concurrent KAD diagnosis

Compared to patients with KS alone, samples from patients with KS and concurrent KAD (KICS or MCD and/or PEL) had differential expression of 26 genes; this included increased expression of the cellular genes STC1 (log2FC=2.02, p-adjusted (padj)=0.001), a secreted glycoprotein, and MKI67 (log2FC=1.11, padj=0.02), a common proliferation marker (**Figure 2A, B**). Pathway analyses identified lower expression of genes associated with specific cytokine activity (normalized enrichment score (NES):-1.95, padj=0.012, **Figure 2C, Supplementary Figure 1**), natural killer cell activation (NES:-2.00, padj=0.028), and B-cell proliferation (NES: −1.93, padj=0.045, **Figure 2C, Supplementary Figure 1**) in lesions from patients with KS with concurrent KAD as compared to those with KS alone.

**Figure 2.**
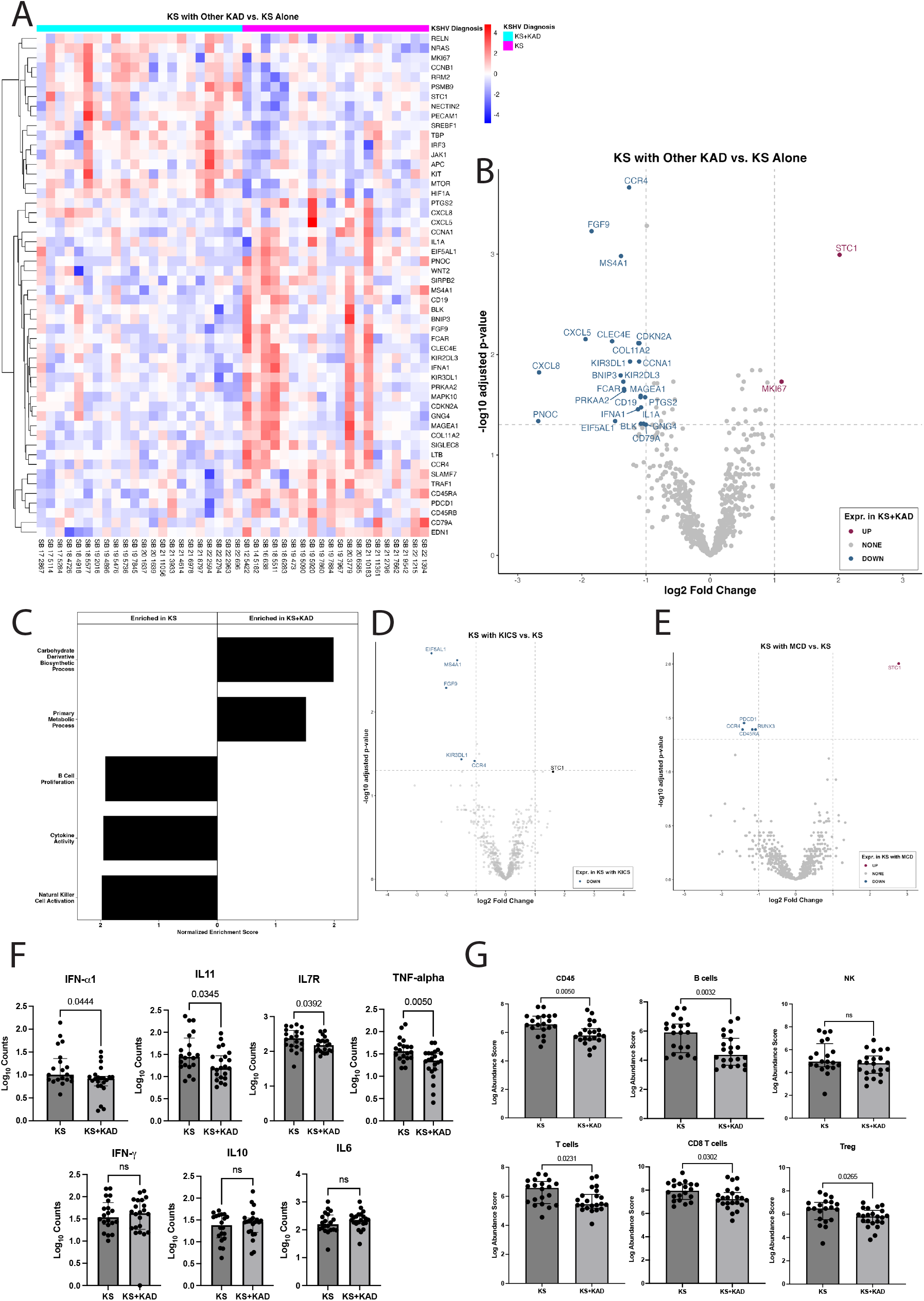
Differential gene expression profiles between KS and specific KAD diagnoses that occur with KS. A. Heat map demonstrating gene expression pattern of KS skin lesions among patients with KS alone and KS with other concurrent KAD B. Volcano plot evaluating differentially expressed genes within KS skin lesion in patients with KS alone (reference) as compared to patients with KS and other concurrent KAD (condition) C. Gene ontology pathway analyses demonstrating differences in pathways between patients with KS and KAD as compared with KS alone D. Volcano plot demonstrating differentially expressed genes within KS skin lesions in 19 patients with KS alone and 13 patients with KICS with KS. E. Volcano plot demonstrating differentially expressed genes within KS skin lesions in 19 patients with KS alone and 6 patients with MCD with KS. F. Selected cytokine gene expression differences between KS lesions from patients with KS alone and those with KS and other KAD (PEL, MCD or KICS). G. Cell deconvolution data demonstrating cell type differences within KS skin lesions between those with KS alone and those with KS and other KAD.

There were differences in gene expression in KS lesions by specific KAD diagnoses as compared to lesions from patients with KS alone. As 13 participants with KICS and KS formed the most abundant group of those with KAD and KS, analyses of their KS lesions as compared to those with KS alone had decreased expression of *CCR4* (log2FC= −1.1, padj= 0.04), *MS4A1* (log2FC= −1.6, padj= 0.002), KIR3DL1 (log2FC= −1.5, padj= 0.04), which are genes associated with immune cell activity. There was also decreased expression of fibroblast growth factor 9 (*FGF9*) (log2FC= −2.1, padj= 0.005), and eukaryotic translation elongation factor 5 (*EIF5AL1*) (log2FC= −2.5, padj= 0.002, **Figure 2D**).

Among 6 participants with KS and MCD compared to KS alone, STC1 was the only differentially expressed gene that was increased in the KS and MCD group (log2FC= 2.8, padj= 0.01, **Figure 2E**). In KS lesions with MCD, there was a notable decrease in *RUNX3* ((log2FC= −1.1, padj= 0.04), PDCD1 (log2FC=-1.4, padj= 0.04), involved in T cell regulation and differentiation, *CCR4* (log2FC= −1.4, padj= 0.04), involved in immune function, and *CD45RA* (log2FC= −1.2, padj= 0.04), involved in lymphocyte regulation and a suppressor of JAK kinases. There were no differentially expressed genes between KS lesions from those with KS alone as compared to those with KS with PEL alone or in PEL with MCD.

Despite the pathway analyses demonstrating decreased cytokine activity among KS lesions from patients with concurrent KAD, individual cytokine levels of interest were evaluated between KS and KS+KAD. Prior studies from our group have demonstrated that in KS+KAD such as KS and MCD or KS and PEL, there are higher levels of IL-6, IL-10, IFN-gamma, TNF-alpha cytokine levels in the circulation during periods of active disease(15, 43). There were higher levels of IFN-alpha, IL-11, IL7R and TNF-alpha in KS lesions from individuals with KS alone as compared to KS+KAD **(Figure 2F**). Notably, when comparing cytokine expression of other cytokines of interest within KS lesions, IL-6, IL-10 and IFN-gamma were not different between KS with or without KAD **(Figure 2F**).

Cellular deconvolution of the nCounter samples highlighted a relative decrease in CD45+ and B cells in specimens from patients with KS and concurrent KAD, particularly in MCD and KICS compared to KS alone **(Figure 2G)**. This may be an expected finding as participants with KS and concurrent KAD may have received prior rituximab-based therapy targeting CD20+ B-cells. T-cells, specifically CD8+ T-cells and Treg expression, were higher in KS lesions from patients with KS alone as compared to those with KS+KAD **(Figure 2G)**.

### HIV viremia and KSHV gene expression levels reveal distinct immune profiles in KS lesions

Seventy-four percent of patients had undetectable HIV viral load (<20 copies/ml) in the blood. In those with detectable HIV levels as compared with well-controlled HIV, KS lesions had higher expression of interferon-stimulated genes (ISGs), such as *OAS1, OAS3*, and *IFIT1*; consistent with heightened immune activation **(Figure 3A)**. Notably, a lytic gene K4, which encodes a viral monocyte inflammatory protein II, was elevated in patients with detectable HIV as compared to those with undetectable HIV (log2FC=2.06, padj=0.048). Based on the median total KSHV ncounter reads, samples were divided into those with high versus low KSHV gene expression. In those with a high KSHV gene expression relative to the median across all samples, there was increased expression of genes *TNFRSF4* (log2FC=1.4, padj=0.001), associated with NF-kappa B activation, BCL6B (log2FC=1.08, padj=0.006), STC1 (log2FC=1.6, padj=0.02), and *MKI67*(log2FC=1.1, padj=0.02, **Figure 3B)**. There was no significant difference in the gene expression profiles of KS lesions when stratifying CD4+ T cell count by 200 cells/µl **(Supplementary Figure 2)**. The median expression of latent genes was higher than the median of lytic gene expression in KS lesions, both among patients with KS alone and in patients with KS+KAD **(Figure 3C)**. This observation of increased latent expression was also noted in lesions from patients with high CD4+ T cell count (≥200 cells/µl) and low CD4 T cell count (<200 cells/µl, **Figure 3D)**. Higher levels of the individual latent genes, K12, K13, ORF72 and LANA, were noted in KS lesions from patients with KS alone and those with KS+KAD **(Figure 3E and 3F)**.

**Figure 3.**
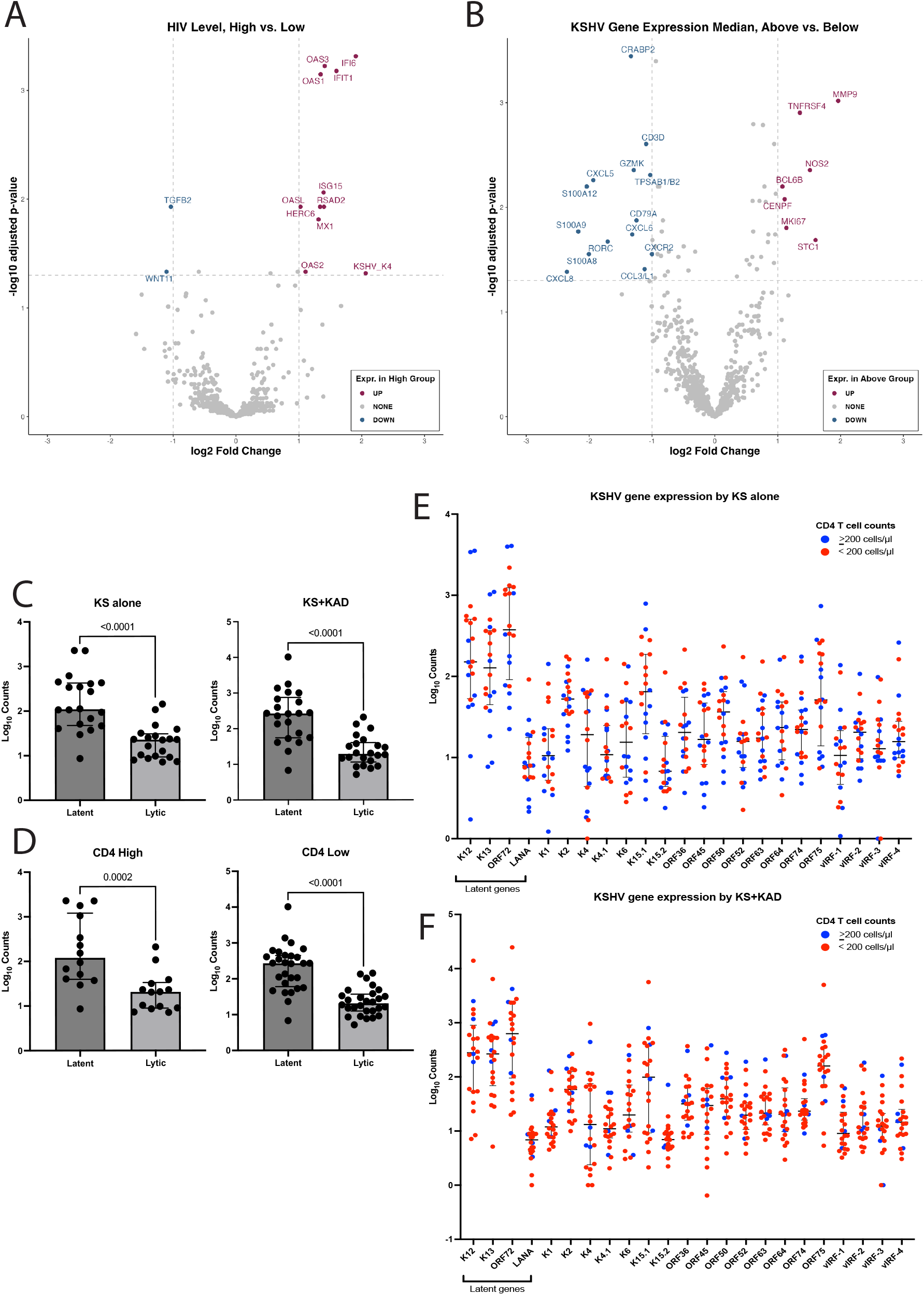
Evaluating gene expression profile differences in KS lesions based on other patient characteristics. A. Volcano plot demonstrating differentially expressed genes within KS skin lesions by HIV viral load (High(>20 copies/ml) vs. Low (≤20 copies/ml)) B. Volcano plot demonstrating differentially expressed genes within KS skin lesions by median of the total KSHV nCounter gene expression profile C. Median expression of latent genes vs. lytic genes in KS lesions among those with KS alone and those with KS and other KAD. D. Median expression of latent genes vs. lytic genes in KS lesions among those with CD4+ T cell High (>200 cells/µl) vs. Low (<200 cells/µl) E. KSHV lytic and latent gene expression levels in KS lesions among patients with KS alone (Blue and Red circles indicate whether lesions were from patients with CD4+ T cell High (>200 cells/µl) vs. Low (<200 cells/µl), respectively. F. KSHV lytic and latent gene expression levels in KS lesions among patients with KS and other KAD (Blue and Red circles indicate whether lesions were from patients with CD4+ T cell High (>200 cells/µl) vs. Low (<200 cells/µl), respectively.

### Transcriptomic factors associated with prognosis

Given differences in survival associated with the presence of KS and other KAD, we investigated whether genes in the nCounter panel were predictive of survival among all patients in the cohort. These analyses were exploratory and were not adjusted for multiple comparisons. Survival analysis demonstrated that high expression of various genes of possible prognostic value **(Supplementary Figure 3)**. High expression of, *TAP1*, involved in MHC class 1 activity, *EZH2*, involved in histone methylation were associated with worse outcomes. Other genes associated with worse outcomes in the cohort included *TNFRSF10D, VEGFC, APOL6*, and *CXCL11*, which are genes associated with chemokine activity and proangiogenic roles. Conversely, *CD2, CD5, CCR4, CTLA4, MS4A2*, and *HLA-A* expression within the KS lesions were noted to have an improved outcome. Many of these genes were linked with T-cell activity, antigen presentation, and immune-related signaling.

### DSP profiling demonstrates differential gene expression in the tumor microenvironment

Digital spatial profiling was performed on 4 KS samples of participants with KS and concurrent KAD: 2 patients had KS and KICS and 2 patients had KS and MCD. The regions of interest (ROI) and areas of interest (AOI) that were indicative of LANA-1+ areas, vascular and immune markers, are highlighted in **Figure 4A**. Cellular deconvolution of DSP specimens highlighted a marked increase in abundance of lymphatic endothelial cells in LANA-1+ tumor areas as compared to uninvolved LANA-1-vessel and immune AOIs **(Figure 4B)**. Conversely, in LANA-1-vessel and immune AOIs, there was a relative increase in vascular endothelial cells, pericytes and mesenchymal fibroblasts, which was expected compared to tumor ROIs. There was a greater abundance of macrophages and T cells noted in the vessel and immune AOIs as compared to the LANA-1+ tumor areas.

**Figure 4.**
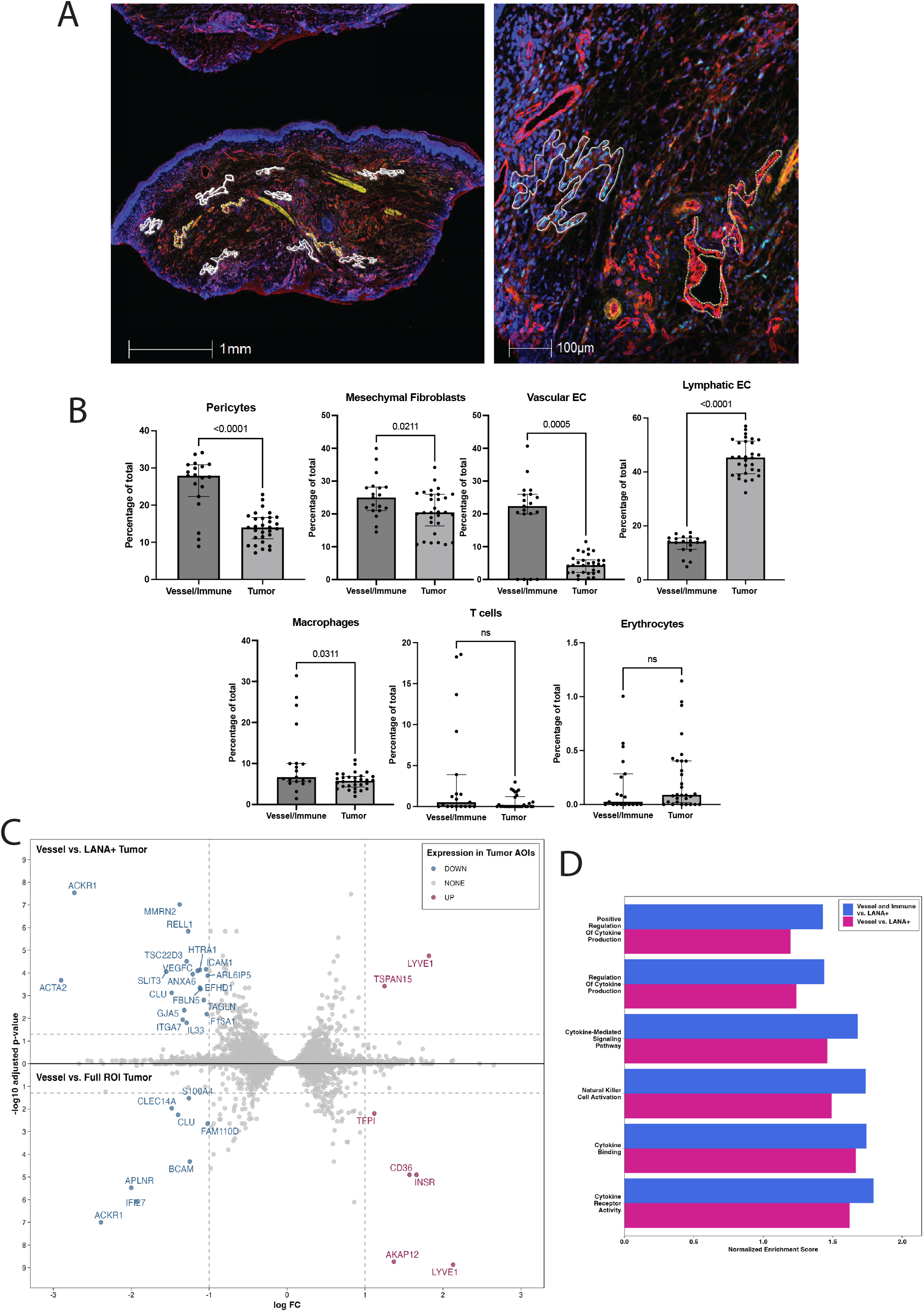
Digital spatial profiling analyses using GeoMx of 4 KS samples from patients with KS and KAD (2 patients with MCD and KS and 2 patients with KICS and KS) A. Representative KS tissue section prepared for GeoMx analysis showing areas of interest (AOIs). Immunofluorescent staining identified distinct cellular compartments: CD45 (yellow) marking hematopoietic cells, CD31 (red) labeling endothelial cells, and LANA-1 (turquoise) highlighting nuclei of KSHV-infected cells. The inset (right) illustrates a KSHV-infected tumor AOI (outlined in solid white) adjacent to an uninfected vessel AOI (outlined in dashed yellow). B. Cell type deconvolution analyses from GeoMx showing increased abundance of lymphatic endothelial cells in tumor areas (LANA-1+) vs vessel and immune areas. C. Double volcano plot showing increased expression of LYVE1 and TSPAN15 in LANA-1+AOIs within tumor AOIs D. Gene ontology pathway analysis demonstrating enrichment of cytokine pathways in vessel and immune areas vs. LANA-1+ areas and in vessel vs. LANA-1+ areas

Analysis of LANA-1+ AOIs identified increased expression of *TSPAN15* (log2FC=1.3, padj=0.0004) and *LYVE1* (log2FC= 1.8, padj= 1.8e-05), both genes are important for oncogenesis and lymphangiogenesis. In these LANA-1+ areas, there were also decreased *ICAM 1* expression(log2FC=-1.04, padj= 6.9e-05), associated with leukocyte chemotaxis and immune cell recognition **(Figure 4C)**.

GSEA analysis of DSP samples highlighted increased enrichment of pathways associated with cytokine-mediated signaling pathway (Normalized Enrichment score (NES)=1.5, padj=0.02) and cytokine binding (NES=1.7, padj=0.03) in the LANA-1 negative AOIs compared to the LANA-1+ AOIs **(Figure 4D)**, suggesting that gene expression associated with cytokine expression is notable in LANA-negative areas in KS specimens. Gene expression of LANA-1+ AOIs was subsequently evaluated in comparison to LANA-1 negative AOIs (immune cell areas and/or vessel areas). Immune cell AOIs had increased cytokine pathway enrichment compared to LANA-1+ AOIs but similar levels compared to those of vessel AOIs such as cytokine-mediated signaling (NES=1.7, padj=2.0e-04), natural killer cell activation (NES=1.7, padj=2.3e-2), regulation of cytokine production (NES=1.4, padj=9.0e-03), cytokine binding (NES=1.7, padj=8.5e-03), cytokine receptor activity (NES=1.8, padj=1.8e-02), and positive regulation of cytokine production (NES=1.4, padj=4.1e-2).

## Discussion

Our study provides novel insights into the heterogenous transcriptomic landscape of KS in PWH, accounting for clinical characteristics, with a particular focus on KS when it occurs with concurrent KADs. Patients in these analyses had well-controlled HIV and 67% of patients had CD4+ T-cell counts <200 cells/µl. CD4+ T cell count levels in patients did not account for gene expression changes in KS lesions. Whereas the presence of a concurrent KAD diagnosis, such as KICS, PEL and/or MCD, was associated with distinct markers of immune dysregulation within KS lesions. We identified the presence of increased *STC1* and *MKI67* in lesions from patients with KS and other KAD and noted that STC1 expression was notably higher in those with KS and MCD than in those with KS alone. Furthermore, from cell deconvolution of nCounter samples, we identified a decrease in B-cell abundance in patients with KS and KAD compared with those with KS alone. DSP analysis highlighted increased *LYVE1* expression, which appeared to correspond with increased lymphatic endothelial cell abundance, in LANA-1+ AOIs in 4 KS samples with concurrent KAD.

Our study identified increased expression of *STC1* expression in tissue samples from patients with KS and concurrent KAD. STC1 expression is a useful prognostic biomarker in many other cancers, including renal clear cell (44), leukemia (45), gastric cancer (46), and hepatocellular carcinoma (47). One study linked the STC1 glycoprotein to breast cancer metastases via the EGFR pathway and the downstream MAPK/ERK signaling, which ultimately contributes to cell cycle progression (48). This effect of *STC1* was thought to be mediated by increased S100A4 expression (48). However, in our analysis,we did not observe a similar association between S100A4 expression and increased STC1 expression. From a prior study published by our group, *STC1* is pro-angiogenic and has also been associated with promoting tubule formation and lymphatic endothelial migration (35, 48)(21). The apparent increased expression of *STC1* within KS among patients with other concurrent KAD suggests that the tumor microenvironment in conjunction with systemic inflammation, and may contribute to the dissemination of advanced stage KS, which is observed in patients with KICS, MCD and/or PEL(43).

Given that prior studies have demonstrated increased inflammatory profiles in the circulation of patients with KAD, it was surprising that the nCounter analyses did not highlight increased expression of pro-inflammatory cytokines within KS lesions of patients with KS and KAD (13, 14, 24, 29, 43). Both nCounter and pathway analyses revealed reduced expression of chemokines, such as CXCL5 and CXCL8, which function as leukocyte attractants, in lesions from patients with KS and concurrent KAD compared with those with KS alone. Higher interferon-alpha 1 expression was also observed in KS alone compared with patients with KS and KICS, indicating greater immunomodulation and an antiviral response to KSHV infection. The lower expression of these chemokine and cytokine-related genes in KS lesions from patients with KS and concurrent KAD may be due to the highly immunosuppressive state of these individuals, who had a median CD4 T cell count of 84 cells/µl. In assessing other specific cytokines, we found no difference in IL-6 and IL-10 expression levels in KS lesions between groups with KS alone and those with KS and concurrent KAD, consistent with our prior study using bulk RNA-sequencing of both skin and GI KS lesions. In the case of MCD, there is evidence that the increased cytokines stem directly or indirectly from KSHV+ plasmablasts in lymph nodes or other lymphoid organs(49, 50). However, the source of the increased cytokines in KICS remains unclear, and the results here suggest that they may stem from sources other than cutaneous.

Although there was no overlap in specific genes between the nCounter and DSP platforms, pathways associated with cytokine regulation and natural killer cell activation were similar in both analyses. These pathways were higher in lesions among patients with KS and KAD compared to those with KS alone and in the DSP platform in 4 lesions from patients with KS and KAD, pathways associated with cytokine production and regulation were higher in immune and/or vessel AOIs compared to LANA+ AOIs. In the DSP platform, we identified increased *LYVE1* gene expression in LANA-1 AOIs, which encodes a type I transmembrane protein, which is a major component of the extracellular matrix, and is a marker of lymphatic endothelial cells. This finding correlated with increased abundance of lymphatic endothelial cells and decreased abundance of vascular endothelial cell populations in LANA-1 AOIs as compared to immune and vessel areas in these patients with KS and KAD. Prior research has indicated increased co-expression of lymphatic markers, such as LYVE-1^+^, in early and late-stage KS using immunohistochemistry, and this correlates with LANA-1^+^ staining(51). TSPAN15, also known as tetraspanin-15, is part of a family of genes implicated in cell adhesion and migration and associated with cancer progression in several cancer cell lines (52). Though its association and role in KS and KSHV infection is unknown, it has been shown to promote the proliferation of hepatocellular carcinoma cells through activation of ERK1/2 signaling(53). Pathway analyses of the DSP identified genes associated with cytokine production and regulation that were higher in the vessel and immune regions, suggesting that cytokine dysregulation may occur in the tumor microenvironment proximal to LANA-1+ regions.

A preliminary analysis of genes associated with KS survival was explored using the nCounter panel. Increased VEGFC expression was also associated with poor outcomes in KS in this cohort(26), as expected, given that aberrant angiogenesis is a hallmark of KS. However, prior studies of anti-angiogenic targeted approaches have not led to robust KS responses(54, 55), and it is unclear whether targeting specific components of this pathway may improve survival outcomes for patients with KS. In these analyses, TAP1 gene, which has several functions associated with immune response and antigen presentation, was associated with worse outcomes. Increased expression of TAP1 was related to poor prognosis in uveal melanoma(56) and clear cell renal cell carcinoma(57). TAP1 protein overexpression was also noted in aggressive breast cancer tissue(58). Another study has shown that increased TAP1 expression in certain cancers conferred a better response to immunotherapy(59). Conversely, genes associated with improved survival were linked to T-cell regulation, antigen presentation, and immune-related signaling. As a tumor that emerges in the context of immune dysregulation and immunosuppression, factors that restore the T-cell response against KSHV infection may improve outcomes for patients with KS.

There are important limitations to consider in this study. First, the retrospective design and lack of normal tissue for comparison prevents us from determining whether the observed gene expression patterns are significant beyond the context of disease. Secondly, the heterogeneity of the study population with regard to the HIV characteristics and prior KS and KAD-directed therapies may have also influenced these findings. Despite these limitations, the study was uniquely valuable in its use of multiple analytic modalities to comprehensively characterize the KS tumor microenvironment in a well-annotated diverse cohort of PWH and KS. The majority of our patients had well-controlled HIV, underscoring the relevance of our findings in the modern clinical landscape. Importantly this cohort included a substantial proportion of individuals with concurrent KADs such as KICS, PEL, and MCD, which are often underdiagnosed, strengthening the clinical relevance of the disease-specific transcriptional programs observed. he use of archival FFPE samples represents an additional strength, as it enables broader real-world applicability and avoids the need for fresh tissue required for bulk RNA sequencing. Finally, integrating nCounter profiling with digital spatial transcriptomics and clinical annotation provided a more holistic perspective on the KS microenvironment.

Although exploratory, our findings suggest that lesion-level molecular phenotyping may eventually complement clinical characteristics in understanding disease heterogeneity in KS. If validated in larger and prospective cohorts, selected markers and spatial features could help refine underlying biological subtypes of KS, identify patients whose lesions are impacted by concurrent KAD diagnoses, or guide enrichment strategies for future therapeutic trials. Future studies should further dissect the paracrine interactions between LANA-1+ tumor regions and adjacent tissues and define how circulating inflammatory signatures interface with local transcriptional programs in patients with KS and concurrent KAD.

In conclusion, these analyses of archival KS tissues using two analytic modalities demonstrated heterogeneous gene expression profiles associated with specific patient characteristics. We observed differentially expressed genes in skin KS samples when stratifying by the presence of concurrent KAD (KICS, MCD and/or PEL). Furthermore, on evaluation of a small subset of KS lesions using spatial transcriptomic technology, we identified genes associated with cell signaling and migration that were upregulated in LANA-1 positive areas as compared with LANA-1 negative areas within the KS tissue specimen.

## Data Availability

All data produced in the present study are available upon reasonable request to the authors

## Prior presentation and publication disclaimer

A part of this study was presented at the American Society of Clinical Oncology (ASCO) 2025, Chicago, IL and the International Conference on Malignancy in HIV 2024, Bethesda, MD. Data from this material should not be interpreted as representing the viewpoint of the U.S. Department of Health and Human Services, the National Institutes of Health, or the National Cancer Institute.

## Funding statement

This work was supported in part, by the Intramural Program of the National Cancer Institute, National Institutes of Health and in part with federal funds from the National Cancer Institute, National Institutes of Health, under contract (75N91019D00024, HHSN261200800001E, ZIA BC 011954, ZIA BC 010888). The contributions of the NIH author(s) were made as part of their official duties as NIH federal employees, are in compliance with agency policy requirements, and are considered Works of the United States Government. However, the findings and conclusions presented in this paper are those of the author(s) and do not necessarily reflect the views of the NIH or the U.S. Department of Health and Human Services.

## Acknowledgements

We sincerely thank the individuals who volunteered for this study and their families for providing their time, energy and effort to contribute to scientific and clinical advancement for these rare conditions. We thank the nurses, physicians, the participant care and clinical research support staff at the National Cancer Institute and the NIH Clinical Center, and the physicians who referred us participants.

## COI disclosures

R. Yarchoan, R Ramaswami and K Lurain report receiving research support from Celgene (now Bristol Myers Squibb), CTI BioPharma (a Sobi A.B. Company), PDS Biotech, and Janssen Pharmaceuticals through CRADAs with the NCI. R. Yarchoan, R Ramaswami and K Lurain also reports receiving drugs for clinical trials from Merck, EMD-Serano, and Eli Lilly and preclinical material from Lentigen Technology through CRADAs or MTAs with the NCI. T.S. Uldrick reports receiving other commercial research support from Roche through a CTA with Fred Hutchinson Cancer Research Center and is now employed by Regeneron Pharmaceuticals. R. Yarchoan is a co-inventor on US Patent 10,001,483 entitled “Methods for the treatment of Kaposi’s sarcoma or KSHV-induced lymphoma using immunomodulatory compounds, and uses of biomarkers.” R. Yarchoan is also a coinventor on patents on a peptide vaccine for HIV and on the treatment of Kaposi sarcoma with IL12, and an immediate family member of R. Yarchoan is a co-inventor on patents related to internalization of target receptors, on KSHV viral IL-6, and on the use of calreticulin and calreticulin fragments to inhibit angiogenesis. All rights, title, and interest to these patents have been or should by law be assigned to the U.S. Department of Health and Human Services; the government conveys a portion of the royalties it receives to its employee inventors under the Federal Technology Transfer Act of 1986 (P.L. 99-502). No potential conflicts of interest were disclosed by the other authors.

## Notes

### Clinical Trial

NCT00006518, NCT03300830

